# Covid-19 affects taste independently of smell: results from a combined chemosensory home test and online survey from a global cohort (N=10,953)

**DOI:** 10.1101/2023.01.16.23284630

**Authors:** Ha Nguyen, Javier Albayay, Richard Höchenberger, Surabhi Bhutani, Sanne Boesveldt, Niko A. Busch, Ilja Croijmans, Keiland W. Cooper, Jasper H. B. de Groot, Michael C. Farruggia, Alexander W. Fjaeldstad, John E. Hayes, Thomas Hummel, Paule V. Joseph, Tatiana K. Laktionova, Thierry Thomas-Danguin, Maria G. Veldhuizen, Vera V. Voznessenskaya, Valentina Parma, M. Yanina Pepino, Kathrin Ohla

## Abstract

People often confuse smell loss with taste loss, so it is unclear how much gustatory function is reduced in patients self-reporting taste loss. Our pre-registered cross-sectional study design included an online survey in 12 languages with instructions for self-administering chemosensory tests with ten household items. Between June 2020 and March 2021, 10,953 individuals participated. Of these, 3,356 self-reported a positive and 602 a negative COVID-19 diagnosis (COVID+ and COVID-, respectively); 1,267 were awaiting test results (COVID?). The rest reported no respiratory illness and were grouped by symptoms: sudden smell/taste changes (STC, N=4,445), other symptoms excluding smell or taste loss (OthS, N=832), and no symptoms (NoS, N=416). Taste, smell, and oral irritation intensities and self-assessed abilities were rated on visual analog scales. Compared to the NoS group, COVID+ was associated with a 21% reduction in taste (95% Confidence Interval (CI): 15-28%), 47% in smell (95%-CI: 37-56%), and 17% in oral irritation (95%-CI: 10-25%) intensity. In all groups, perceived intensity of smell (r=0.84), taste (r=0.68), and oral irritation (r=0.37) was correlated. Our findings suggest most reports of taste dysfunction with COVID-19 were genuine and not due to misinterpreting smell loss as taste loss (i.e., a classical taste-flavor confusion). Assessing smell and taste intensity of household items is a promising, cost-effective screening tool that complements self-reports and helps to disentangle taste loss from smell loss. However, it does not replace standardized validated psychophysical tests.

## Introduction

The SARS-CoV-2 virus had a major impact on all three chemical senses involved in flavor perception: smell, taste, and chemesthesis. While smell loss became known as the cardinal symptom of COVID-19 and was studied extensively, the veracity of the associations between COVID-19 and taste loss have been called into question (Hannum et al., 2022; Saniasiaya et al., 2021). Unlike the relatively common loss of smell seen with viral infections of the upper respiratory tract (Seiden, 2004), post-viral taste loss was rare prior to the pandemic (Henkin et al., 1975; Pribitkin et al., 2003) presumably due to common semantic confusion between taste, smell, and flavor (Boltong and Keast, 2012). Thus, most patients who present to the clinic with “taste” complaints actually have normal gustatory function, and systematic evaluation of the chemical senses instead indicates the presence of olfactory dysfunction (Pribitkin et al., 2003). Notably, less than 1% of 1,176 patients who presented to a specialized clinic reporting olfactory and/or gustatory dysfunction had actual taste loss, while 32% had smell loss (Pribitkin et al., 2003). That is, discrepancies between patients’ reports and results of direct evaluations are likely due to the common confusion between taste and smell or flavor. Much of what the public describes colloquially as “taste” encompasses not only taste but also retronasal smell (Murphy et al., 1977; Murphy and Cain, 1980) and chemesthesis (Duffy and Hayes, 2020). Because loss of retronasal smell is often mischaracterized as taste loss by both patients and clinicians, it remains unclear to what extent taste and chemesthesis are affected when individuals lose their sense of smell, such as in COVID-19.

While numerous survey-based studies, including our own (Parma et al., 2020; Gerkin et al., 2021), suggest that COVID-19 affects taste function (see (Saniasiaya et al., 2021), for a meta-analysis), studies that used various kinds of taste stimuli (versus self-report) have yielded conflicting results. In studies with self-administered home tests, where participants with COVID-19 prepared four solutions with prototypical tastants and identified the taste quality, taste dysfunction was seen in 42-72% of participants (Petrocelli et al., 2020; Vaira et al., 2020a, 2020c, 2020b). Conversely, studies that used standardized taste tests, such as taste strips (Hintschich et al., 2020; Le Bon et al., 2021a, 2021b) or the Waterless Empirical Taste Test (Cao et al., 2022a) suggest the sense of taste was generally well preserved (Hintschich et al., 2020) or only mildly affected in individuals with acute COVID-19 infection (Haldrup et al., 2020; Le Bon et al., 2021a, 2021b), causing some authors to speculate that reports of taste loss with COVID-19 were probably due to taste/flavor confusion and a loss of retronasal smell (Hintschich et al., 2020). Alternatively, clinic-based assessments with validated tools may have still missed true taste loss if the dysfunction was transient and resolved by the time testing occurred in a clinical setting.

The primary purpose of the present study was to determine to what extent taste function -as well as smell and oral irritation - were affected by COVID-19. To this end, we designed a remote online chemosensory home test that independently captures the intensity of taste, smell, and oral irritation of common household items in parallel with self-assessments of chemosensory ability using a previously published online survey (Parma et al., 2020; Gerkin et al., 2021; Albayay et al., 2022; Ohla et al., 2022; Weir et al., 2022).

First, we hypothesized that individuals with COVID-19 would show greater impaired chemosensation as compared to people with other respiratory illnesses or no symptoms. Specifically, we expected participants in the COVID-19+ group to rate household items as being less intense than those in COVID- and NoSymptoms groups for all three chemosensory modalities. To substantiate this hypothesis, we further explored whether perceived intensities of chemosensory stimuli would be most reduced closer to the onset of COVID-19 rather than later on in the disease (>15 days post symptom onset), as worst chemosensory function had been recorded during the first week of infection (Vaira et al., 2020b).

Second, we hypothesized that the self-reported ability to taste, smell and perceive oral irritation be positively associated with the experienced intensity of tastants, odorants and stimuli irritating the oral mucosa, respectively. As caveats, we anticipated that the association between self-reported ability and perceived intensity would be stronger for odorants than for odorless tastants, because of taste-smell confusion. In other words, we expected some participants to rate the intensity of odors as greater as compared to the intensity of odorless tastants, such as sugar and salt.

Lastly, we explored in a data-driven manner the existence of chemosensory profiles across the studied sample and we characterized a subgroup of individuals with taste/smell confusion.

## Methods

Data were collected using a crowd-sourced, multilingual, online study with a multi-national reach: the GCCR Smell-&-Taste-Check deployed, at the time, in 12 languages (Czech, Dutch, English, French, German, Italian, Japanese, Korean, Portuguese, Russian, Spanish, Turkish). For chemosensory self-assessment, participants were asked to rate their abilities to smell, taste, and perceive oral irritation via the Global Consortium for Chemosensory Research (GCCR) survey (Parma et al., 2020; Gerkin et al., 2021). For empirical chemosensory data, participants rated the intensity of the smell or taste of common household items (e.g., shampoo, fruit juice, sugar, etc.) as appropriate for the stimulus, and the intensity of nasal and oral irritants (e.g., smelling vinegar, tasting mustard). Participants were provided with a list of 75 items that were selected to be culturally diverse. These included 42 odors (including 7 cosmetics/detergents, 22 spices, 13 fruits/vegetables, and 17 other items), 4 tastes (including sweet, sour, bitter, salty), and 8 oral and 4 nasal irritants (Supplementary Table 1). Participants were instructed to select ten items (one per category) available in their household. Self-reported chemosensory abilities and perceived chemosensory intensities of the household items were reported on a 101-point visual analog scale (0-100) anchored with “no sensation” and “very intense” (Parma et al., 2020; Gerkin et al., 2021). Additionally, we collected data on demographics, including age, gender, pregnancy (for women only), education, smoking, socio-economic status (SES), number of social contacts, current symptoms, COVID test status, and medical history.

The study was publicized via the GCCR website (https://gcchemosensr.org) and social and traditional media. All volunteers provided consent and confirmed that they were 18 years old or older. The study procedure generated no dropouts because only complete data sets were recorded in the database. However, participants had the option to skip items, leading to incomplete data for some variables and different sample sizes for the different sensory modalities. The institutional review board of the Faculty of Psychology & Sports Science at the Westfälische Wilhelms-Universität Münster approved the study (no. 2020-27-NB). Additional approval was obtained for the Russian test version by the Bioethics Committee at the A.N. Severtsov Institute of Ecology & Evolution RAS (no. 2020-41-NC). Consent was documented electronically within the online study interface.

### Participants

In total, 10,953 participants completed the survey between June 26, 2020, and March 23, 2021. Of these, 35 were excluded because they provided implausible data (no COVID diagnosis, but COVID onset date). Data from the remaining 10,918 participants are reported here, and their group age and gender information are reported in Figure 1. Of these, 5,225 participants reported a respiratory illness and were subsequently asked about their COVID-19 status. A total of 3,356 reported a positive and 602 a negative COVID-19 test result or clinical diagnosis; these groups were classified as COVID+ and COVID-, respectively. Individuals with a respiratory illness who were awaiting test results were classified as COVID unknown (COVID?; N=1,267). The remaining 5,693 reported no respiratory illness and were, as per survey design, not asked about COVID-19, but only about current symptoms. Based on reported symptoms, we grouped these remaining participants into those reporting sudden smell/taste changes (STC, N=4,445), other symptoms excluding smell or taste loss (OthS, N=832), and no symptoms at all (NoS, N=416). Demographics of the six groups are summarized in Supplementary Table 2.

**Figure 1.**
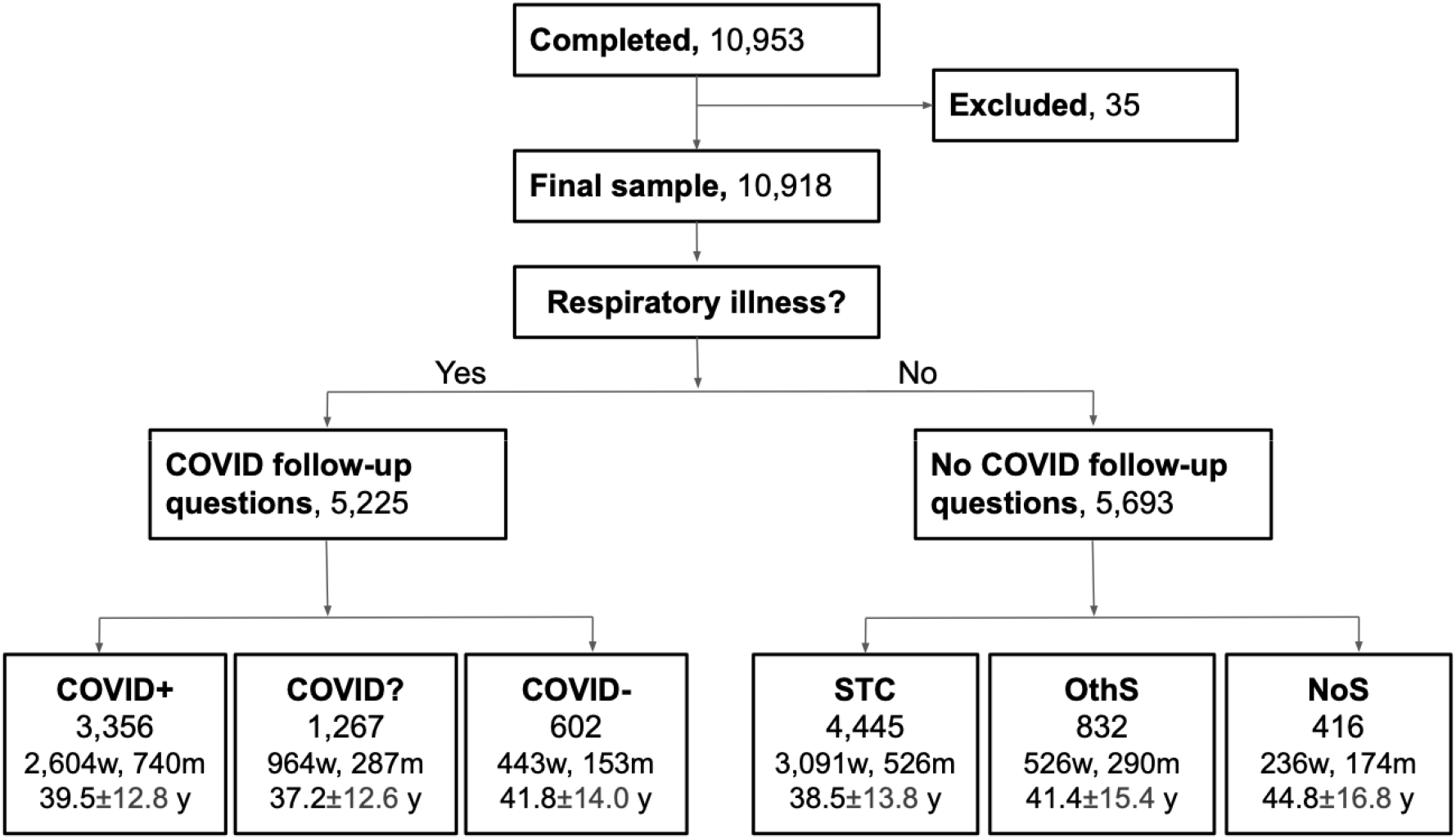
Flow diagram showing the participants included in the study. Exclusions were based on implausible data. Participants were split based on their response (yes/no) to the question asking whether they have a respiratory illness. Only those who responded with “yes” to the respiratory illness question were asked whether they had COVID-19. They were further split according to their report of a positive (COVID+) or negative (COVID-) diagnosis or if they were waiting for a test result and were suspected to have COVID-19 (COVID?). Those who responded “no” to the respiratory illness question were split by the symptoms reported: only smell and/or taste-related changes (STC), any OtherSymptom but smell/taste changes (OthS), or NoSymptoms at all (NoS). Age ± SD in years (y); Gender is reported as women (w) and men (m), remaining participants identified as “other” or did not share.

### Data analyses

Data analyses were performed in R version 4.0.4 (R Core, 2021) using the *tidyverse+* (Wickham et al., 2019), *FactoMineR* (Lê et al., 2008), *cluster* (Maechler et al., 2022), *plotly (Sievert, 2019), car* (Fox and Weisberg, 2019), and multcomp (Hothorn et al., 2008) packages. We computed separate smell, taste, and oral irritation composite scores based on the average of all available perceived intensity ratings by chemosensory modality because participants could opt to smell as many as four household items and taste four different foods. In contrast, they tasted and smelled only one irritating item to minimize carryover of irritation. Missing ratings were not imputed leading to variable sample sizes for different analyses.

To test the primary pre-registered hypothesis (https://osf.io/6bfua) that participants with COVID-19 would exhibit a reduced perceived intensity to taste, smell and irritating stimuli compared to participants without COVID-19 and healthy individuals, participants were grouped according to their COVID-19 status, and data were submitted to separate analyses of variance (ANOVAs). In these analyses, we compared six groups (COVID+, COVID-, COVID?, STC, OthS, NoS) on perceived intensity of the household items and self-reported ability (without specific stimuli) to smell, to taste, and to perceive oral irritation. For the comparisons between ability and intensity for chemesthesis, we used oral irritation ratings as these were included in both the survey and the hometest.

To test the secondary pre-registered hypothesis that self-reported abilities to taste, to smell and to perceive oral irritation were positively associated with the respective perceived intensities of real-life stimuli, we computed Spearman correlation coefficients between the two measures for each sense separately. We then compared coefficients using the *cocor* package in R (Diedenhofen and Musch, 2015) to test the hypothesis that the association between self-reported ability and perceived intensity for smell would be stronger than the association between self-reported ability and perceived intensity for taste and oral irritation.

Additionally, we explored whether perceived chemosensory intensity was influenced by the time passed since the onset of COVID-19 (worst chemosensory function during the first week of infection, we compared with an ANOVA the intensity scores of COVID+ individuals who participated in this study within 0-7 days (N=737), 8-14 days (N=597), or more than 15 days (N=1,831) since the onset of COVID-19 symptom.

An exploratory Multiple Factor Analysis (MFA) using *FactoMineR* was performed to test the rationale of our a priori grouping. We present a map of the six diagnostic groups (see Figure 1) based on the chemosensory profiles from the self-reported abilities and perceived intensities of taste, smell, and oral irritation. Each group is circled by a 95% confidence ellipse generated by virtual panels using Bootstrap techniques, and a graph of a correlation circle of the ratings.

Finally, we used Agglomerative Hierarchical Clustering (AHC) with Ward’s clustering method (Nielsen, 2016) on the self-reported ability and perceived intensity of taste, smell, and oral irritation from the entire sample to differentiate participants solely based on their chemosensory profiles and irrespective of diagnosis in a data-driven manner. This AHC analysis revealed three clusters of participants with distinct chemosensory profiles, one of which confused smell dysfunction for taste dysfunction.

The chemosensory abilities and perceived intensities of the three resulting clusters were compared by one-way ANOVA and Tukey’s HSD test. Additionally, to explore the characteristics of the identified clusters, we analyzed the continuous demographic variables with one-way ANOVA and Tukey’s HSD test and categorical variables with Chi-squared test. The alpha level for all analyses was set a priori at 0.05. Corrections for multiple comparisons were made as necessary. Effect sizes were computed for inferential tests. The data were assessed for the assumptions of the respective statistical tests employed. For details, please refer to the script used for analyses (https://osf.io/6bfua).

## Results

### Reduced perceived intensity of taste, smell, and oral irritation stimuli in COVID-19

Groups reporting confirmed or suspected COVID-19 (COVID+, COVID?, and STC groups) rated the intensity of taste, smell, and oral irritation from foods and household items as being less intense than the COVID-, OthS and NoS groups (Group effect for Smell: F_(5, 10536)_ = 178.8, p < 0.001, η_p_ ^2^ = 0.08, Taste: F_(5, 10541)_ = 70.5, p < 0.001, η_p_ ^2^ = 0.03, and oral irritation: F_(5, 8424)_ = 40.16, p < 0.001, η_p_^2^ = 0.02; Figure 2). Compared to the NoS group, COVID+ exhibited a 21% reduction in taste intensity (95% CI: 15-28%), a 47% reduction in smell intensity (95% CI: 37-56%), and a 17% reduction in oral irritation intensity (95% CI: 10-25%; see Supplementary Table 3 for pairwise tests). To confirm reductions in retronasal smell did not drive reductions in ratings of taste intensity, we re-analyzed the data on perceived taste intensity by selecting stimuli that are purely gustatory in nature (i.e., salt and sugar) and excluding taste stimuli that also had an odor (e.g., lemon juice or coffee beans). The subset of data that included only odorless taste stimuli confirmed the results seen with data from all stimuli (F_(5, 10386)_ = 57.2 p < 0.001, η_p_^2^ = 0.03; Supplementary Figure 1).

**Figure 2.**
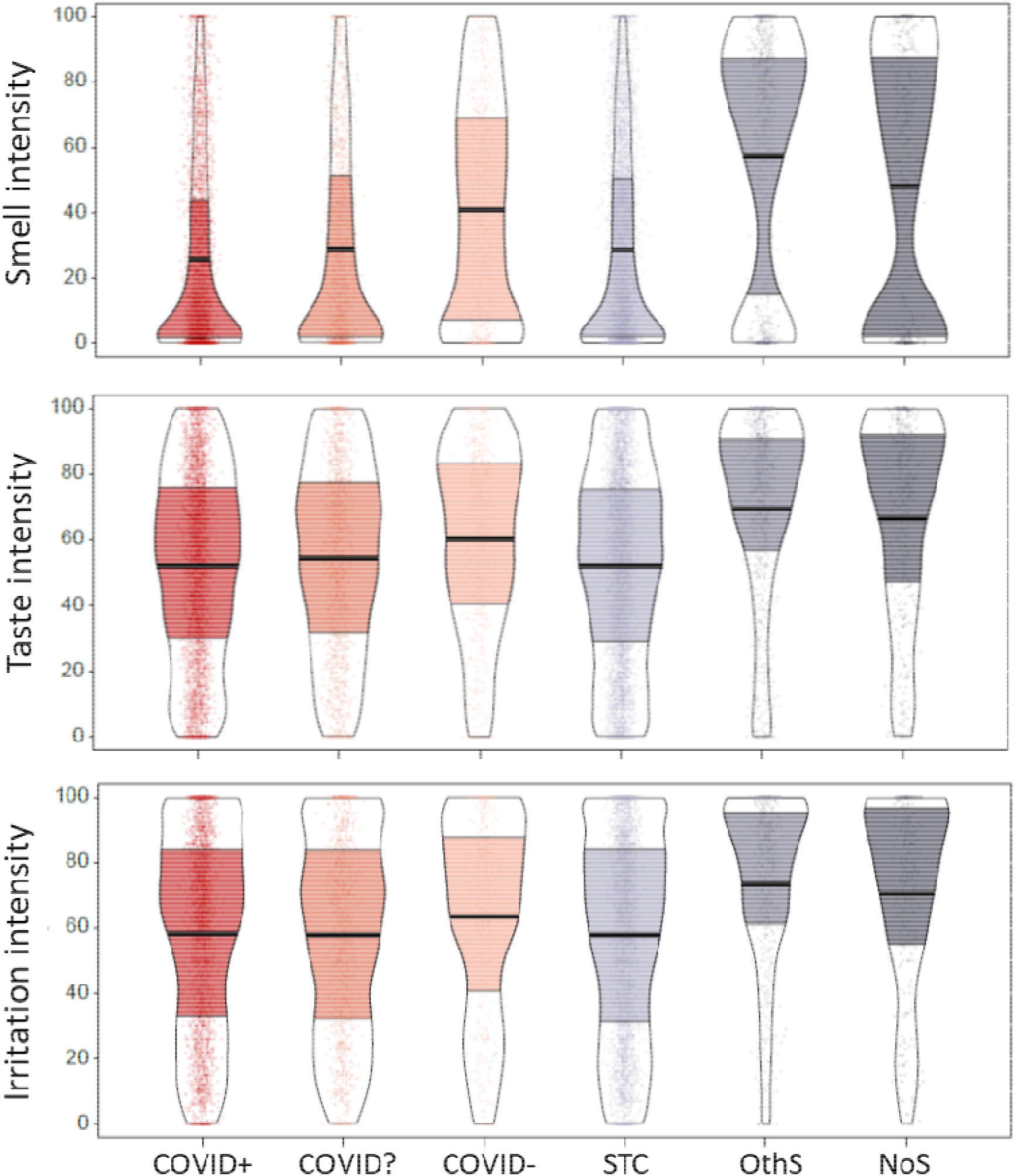
Perceived intensity of taste, smell, and oral irritation when sampling food or household items for six groups of participants. Participants are grouped according to COVID diagnosis or symptoms (from left to right) into COVID positive (COVID+; N=3,275), unknown COVID status (COVID?; N=1,224), and COVID negative (COVID-; N=579), those who reported sudden smell/taste changes (STC; N=4,271), those with other symptoms excluding smell or taste loss (OthS; N=802), and those with no symptoms (NoS; N=396). They rated perceived intensity of smell, taste, and oral irritating stimuli using a visual analog scale (0-100). Points represent individual subject data (jittered horizontally), the center horizontal bars depict the median, the shapes reflect the density of the distribution, and the colored areas show interquartile ranges. For a similar presentation of data for self-reported chemosensory ability, see Supplementary Figure 3.

To test the hypothesis that reductions in the perception of taste, smell, and oral irritation were related to acute COVID-19 infection, we compared the intensity ratings of individuals who completed the survey within the first 7 days (n=737), 8-14 days (n=597), and >15 days (n=1,831) from the onset of their COVID-19 symptoms. Intensities differed between these groups for all three chemosensory modalities (Smell: F_(2,3099)_ = 524.9, p < 0.001, η_p_ ^2^ = 0.25, Taste: F_(2,3091))_ = 153.0, p < 0.001, η_p_ ^2^ = 0.09, and irritation: F_(2,2573)_ = 56.3, p < 0.001, η_p_^2^ = 0.04; Supplementary Figure 2). Post-hoc tests revealed the lowest smell, taste, and oral irritation intensities for those individuals who completed the survey within 7 days, higher intensities for those who completed the survey between day 8 and 14, and the highest intensities were found for those who participated 15 days or more after symptom onset. All pairwise comparisons were significant except oral irritation which did not significantly differ between the first two-time segments (Supplementary Table 4).

### Strong links between chemosensory intensity perception and self-reported ability, but a subset of individuals show taste/smell confusion

We found medium to strong correlations between self-reported chemosensory ability and the rated intensity of the chemosensory stimuli for smell (r=0.84), taste (r=0.68), and oral irritation (r=0.37; all p<0.001; see Supplementary Figure 4), with significantly stronger associations for smell than for the other two chemical senses (p<0.0001).

To corroborate the finding from the bivariate analyses, we also performed a multiple factor analysis (MFA) across all data, finding that self-assessment of chemosensory ability was associated with the respective ratings of chemosensory intensity (Figure 3A). Regardless of the item selected, smell ratings were correlated across all categories (that is, cosmetics, spices, fruits, and other). Similarly, all taste ratings correlated across categories (namely, sweet, sour, salty, and bitter) as well as with ratings of stimuli eliciting oral irritation. The distribution of the a priori identified diagnostic groups suggest that Dimension 1 reflects a spectrum from respiratory illness to respiratory health and Dimension 2 reflects chemosensory function from low to high. The proximity and overlap of the confidence ellipses of the COVID+, COVID?, and STC groups suggests these may belong to one latent group that is characterized by substantial impairment of chemosensory function (Figure 3B).

**Figure 3.**
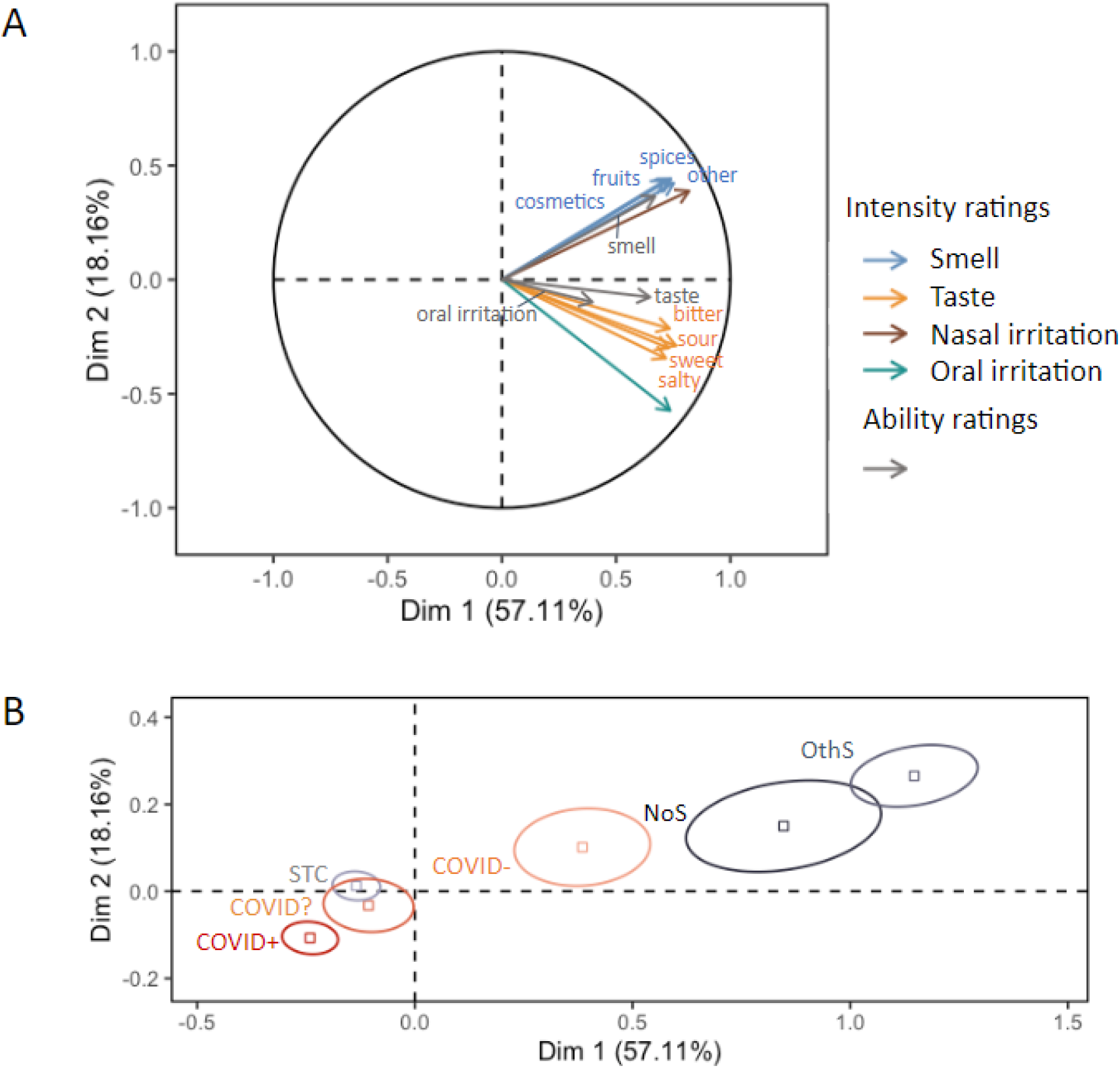
Multiple Factor Analysis (MFA) on self-reported chemosensory abilities and perceived intensities. (A) Correlation circle including all ratings. (B) Map of the six groups (COVID+, N=3,275; COVID-, N=579; COVID?, N=1,224; STC, N=4,271; OthS, N=802; NoS, N=396) with 95% confidence ellipses.

To explore the relative contribution of each chemosense to the integrity of the overall chemosensory function in a data-driven manner, we applied AHC to ratings of chemosensory ability and intensity regardless of diagnosis. As shown in Figure 4, three chemosensory cluster profiles were identified: the “minimally impaired” for all three senses, with good correspondence between ratings of intensity of food and household items and self-report ability to smell/taste/perceive oral irritation (Cluster 1), the “severely impaired” for all three senses, with good correspondence between intensity ratings and self-reported ability (Cluster 2) and the “severely smell impaired” who exhibited, good correspondence between intensity ratings and self-reported ability for smell but not for taste, and to some extent oral irritation. Specifically, respondents self-reported lower ability than intensity of the same items, suggesting that they conflated their reduced ability to smell with a reduced ability to taste and perceive oral irritation rather than a true lack of taste ability, indicating that these participants may have possibly confounded taste and irritation with smell (Cluster 3). The demographics of all three clusters are summarized in Supplementary Table 4.

**Figure 4.**
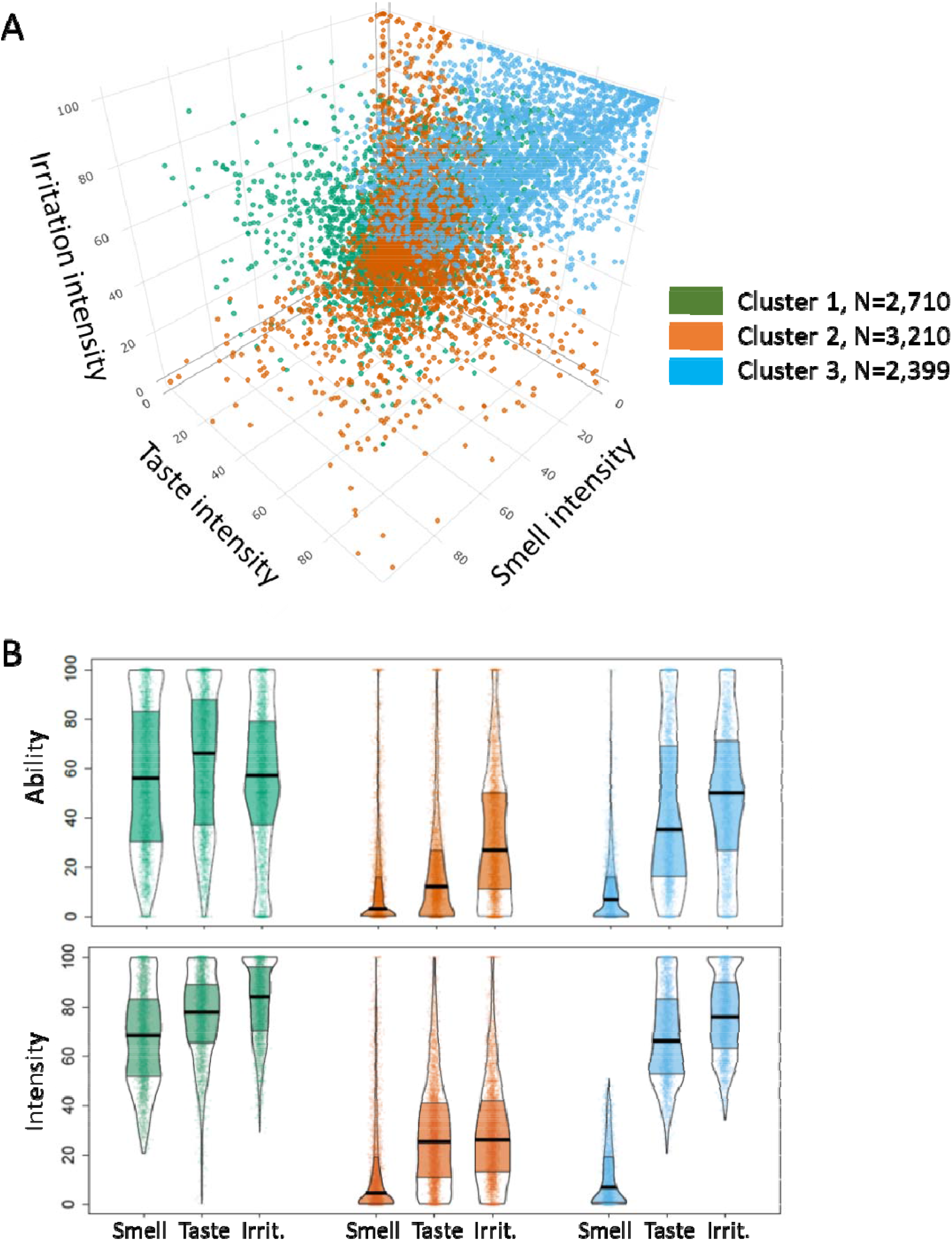
Differences in self-reported abilities and perceived intensities for smell, taste, and oral irritation between three clusters obtained by the Agglomerative Hierarchical Clustering (AHC) on perceived intensities irrespective of reported diagnosis. (A) 3D plot on smell, taste, and oral irritation intensities. Dots represent individual subject data, clusters are color-coded. (B) Self-reported abilities and perceived intensities of foods and items of the three chemosensory modalities of smell, taste, and oral irritation for the three clusters. Points represent individual subject data (jittered horizontally), the center horizontal bars depict the medians, the shapes reflect the density of the distribution, and the colored areas show the interquartile range. Cluster one (green) was minimally impaired, while cluster 2 (orange) was severely impaired for all three chemical senses; cluster 3 (blue) showed severe loss of smell but not of taste or oral irritation.

## Discussion

Taste loss and smell loss have been cardinal self-reported symptoms of infection, particularly with early variants of the SARS-CoV-2 virus (Coelho et al., 2022). With any smell loss coincident to viral illness, patients often present with complaints of “taste loss” that manifest as changes in food flavor, which result from congestion causing an impaired (retronasal) smell. Very early in the pandemic, initial reports of impaired taste were widely assumed to reflect this sort of taste/smell/flavor confusion but growing evidence from patient reports suggest COVID-19 might also impact the sense of taste (unlike the typical cold). Here, we use a large study of 10,953 individuals to demonstrate that COVID-19 not only affects smell but also taste and chemesthesis, at least in individuals who became ill between June 2020 and March 2021. The finding of a reduced perceived taste intensity for pure gustatory stimuli like sugar and salt suggests that COVID-19-associated complaints of taste loss are real and not merely a result of classic taste-smell confusions, in a majority of individuals. A hierarchical cluster analysis corroborated this finding.

Findings from our study confirm and extend results from previous studies on the utility of assessing ratings of perceived intensity for actual chemosensory stimuli to evaluate chemosensory dysfunction (Iravani et al., 2020; Snitz et al., 2022). For smell, the intensity of household items was reduced by 47% in the COVID-19+ group compared to the presumably healthy NoSymptoms group. Extending previous findings, we found that taste intensity and oral irritation intensity when tasting real food items were also markedly reduced by 21% and 17%, respectively, in COVID-19+ participants. Reductions in chemosensory intensity were most profound in those participating during the first two weeks of their COVID-19 illness, thus supporting the interpretation that there is a direct causal relationship between COVID-19 and the broad loss of chemosensation.

Significant progress has been made to understand the cellular and molecular mechanisms of COVID-19-associated smell loss. Findings from postmortem samples of respiratory and olfactory mucosae and whole olfactory bulbs of COVID-19 patients who died a few days after infection with SARS-Cov-2 confirmed previous inferences (Cooper et al., 2020; Khan et al., 2021) that the sustentacular cells, but not the olfactory sensory neurons, are the main target for SARS-CoV-2 (Cooper et al., 2020; Khan et al., 2021). These findings suggest that the sudden anosmia in COVID-19 is caused by olfactory sensory neurons lacking adequate support from sustentacular cells (Finlay et al., 2022; Butowt et al., 2023). Although less is known about the underlying mechanisms of impairment of taste and oral irritation in COVID-19, it is clear that SARS-CoV-2 infects the oral cavity (Huang et al., 2021). The virus is found in saliva, and human taste cells express essential mechanisms of entry of the virus into the host cell, including the angiotensin-converting enzyme 2 (ACE2) and the transmembrane protease serine 2 (TMPRSS2) (Sakaguchi et al., 2020; Doyle et al., 2021). Interestingly, it has been shown that patients’ self-reports of smell/taste loss are positively associated with salivary viral load (Huang et al., 2021). Furthermore, taste function could be affected by the high levels of the pro-inflammatory cytokines -TNF-α, IFN-γ, and IL-6 observed in serum of COVID-19 patients which can impair stem cell function, and hence, taste bud cell renewal (Doyle et al., 2021). Another complementary mechanism contributing to decreased taste perception in individuals with anosmia could be the abolition of the enhancement of taste by smell that is assumed in healthy individuals (Hintschich et al., 2020; Ai and Han, 2022)

Subjective ratings of olfactory function are considered by some to be unreliable and inaccurate because respondents are prone to under-or over-reporting biases or may use response scales idiosyncratically. However, subjective olfactory ratings may still be accurate reflections of ability when certain conditions are met. For example, olfactory ratings of people with severe hyposmia or anosmia have an accuracy rate of 70-80% (Lotsch and Hummel, 2020). In other words, individuals may be unaware of loss until formally examined (Landis et al., 2003), but those who do report chemosensory impairment have a high likelihood of this being a genuine symptom. That is, the relationship is asymmetrical - true loss may not be noticed until testing, but self-reports of loss are typically accurate. Furthermore, questions about smell or taste are frequently ambiguous; a classic example is asking respondents to comment on their sense of taste without specifying that the question is specific about basic taste qualities such as sweetness and not flavor (taste/smell confusion) (Rozin, 1982; Gerkin et al., 2021; Weir et al., 2022). Furthermore, the scale associated with a question may be insufficiently granular (Cao et al., 2022b). For example, drawing from simple surveys used to assess sino-nasal or oral symptoms prior to and during the early stages of the COVID-19 pandemic, “loss of taste and smell” was frequently presented as a single symptom to be reported on a binary (yes/no) question in clinical tests like the SNOT-22 (Kennedy et al., 2013)]. Questions about smell and taste experiences when using dichotomous questions versus visual analog scales, provide different information of an individual’s ability to experience smell, taste, and chemesthesis during and after COVID-19 or other respiratory illnesses (Gerkin et al., 2021). The lack of granularity from assessments that rely on identification rather than intensity, for example “Do you taste anything?” followed by “Do you recognize the taste?” after tasting different foods, might also be the reason why recent home taste tests failed to be associated with standardized tests in the clinic - although this approach produced reliable results for olfactory function (Li et al., 2022). One of the current work’s strengths is the unambiguous description of the sensory experience to be measured (taste vs. smell), as well as the use of continuous visual analog scales for both ability and perceived *intensity* of samples chemosensory stimuli, which we found to be highly correlated (0.84), supporting the notion that self-reports of olfactory ability can be accurate and informative (Li et al., 2022).

Another strength of our crowdsourced study is the large sample size and sizable global recruitment spread as well as the combination of self-administered and self-assessments of smell, taste, and oral irritation intensity measures. Our data also includes non-COVID control participants, unlike other studies using home tests that included only participants with COVID. Because our questionnaire and home tests were administered in several countries and languages, we acknowledge that we cannot exclude biases or effects owing to language differences (Weir et al., 2022). However, we tried to minimize these effects through a standardized translation protocol (Brislin, 1970; Moshontz et al., 2018). Also, our study population is a self-selected convenience sample that may be biased towards inclusion of participants with an increased interest in smell and taste and/or their disturbances.

With the SARS-CoV-2 pandemic, the number of people affected by chemosensory impairment has been steadily increasing, and it will take years to reveal the extent of long-term and chronic chemosensory damage in the population. Notably, our study demonstrates genuine taste dysfunction in participants with COVID-19. Basic and translational research is needed to further our understanding of the underlying mechanisms of post-viral taste and smell impairment and recovery. From a clinical perspective, inexpensive taste and smell tests using household items - even if only collected at home - have utility in screening for chemosensory dysfunction in COVID-19 and may also have similar applicability for aging, and neurodegenerative diseases. Such approaches can support the clinical diagnosis with standardized psychophysical tests and the selection of appropriate treatment options as well as the monitoring of recovery of chemosensory function.

## Data Availability

Data and code will be made available in the project Github repository after publication with Creative Commons Attribution-NonCommercial-NoDerivatives 4.0 International Public License (CC BY-NC-ND 4.0; https://creativecommons.org/licenses/by-nc-nd/4.0/).

## Acknowledgement

The authors wish to thank all study participants and all members of the Global Consortium for Chemosensory Research (GCCR) for help with translations and survey design.

## Author contribution

All authors contributed to multiple aspects of the research, including conceptualization of various aspects of the design and analysis, data collection and interpretation, and critical reviewing. H.N. and J.A. led the analytical efforts and are co-first authors; R.H. led the design of the online survey and implemented the online study; V.P., M.Y.P and K.O. led the design of the survey, supervised all aspects of the project, led the drafting and revision of the manuscript and are co-last authors; all other authors are alphabetically listed to indicate equal contributions.

## Supplementary material

## Supplementary Figures

**Supplementary Figure 1.**
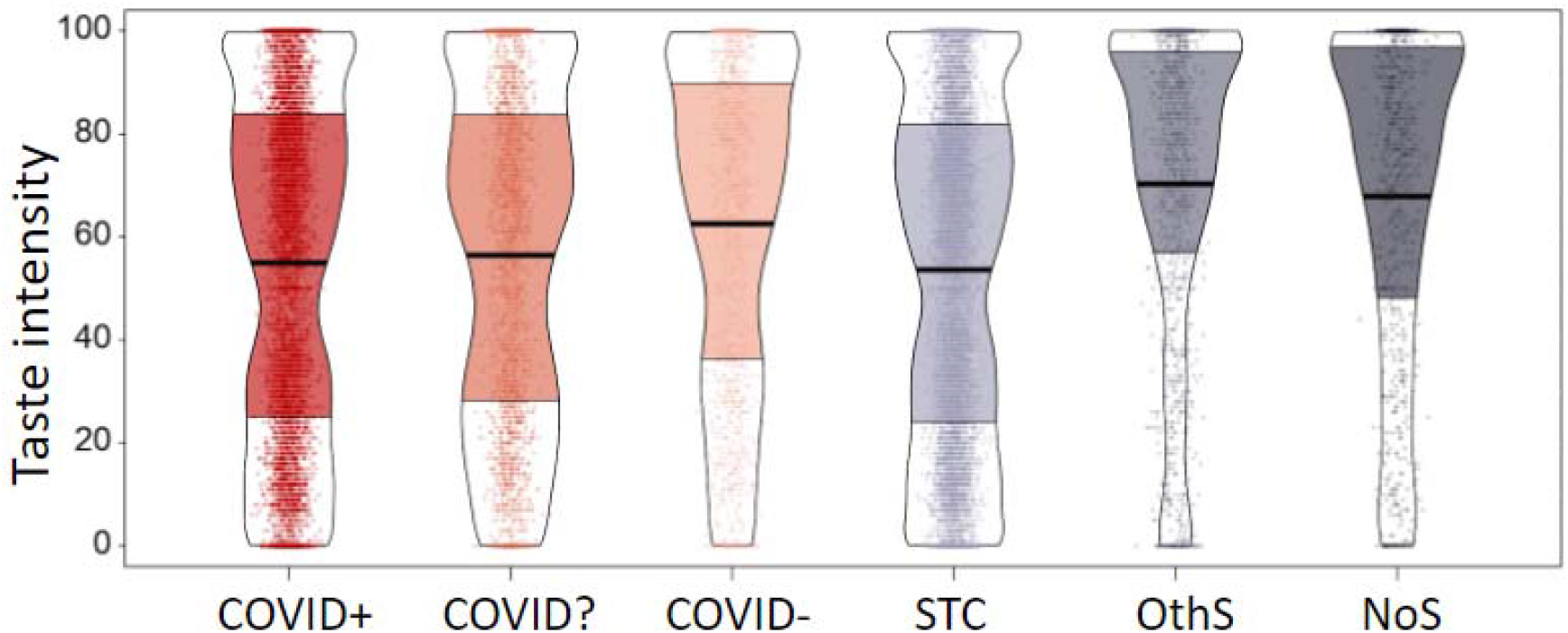
Perceived intensity of pure taste items (sugar and salt) for six groups. Participants are grouped according to COVID diagnosis or symptoms (from left to right) into COVID positive (COVID+; N=3,275),, and COVID negative (COVID-; N=579), those who reported sudden smell/taste changes (STC; N=4,271), those with other symptoms excluding smell or taste loss (OthS; N=802), and those with no symptoms (NoS; N=396). They rated taste intensity using a visual analog scale (0-100). Points represent individual subject data (jittered horizontally), the center horizontal bars depict the median, the shapes reflect the density of the distribution, and the colored areas show interquartile ranges.

**Supplementary Figure 2.**
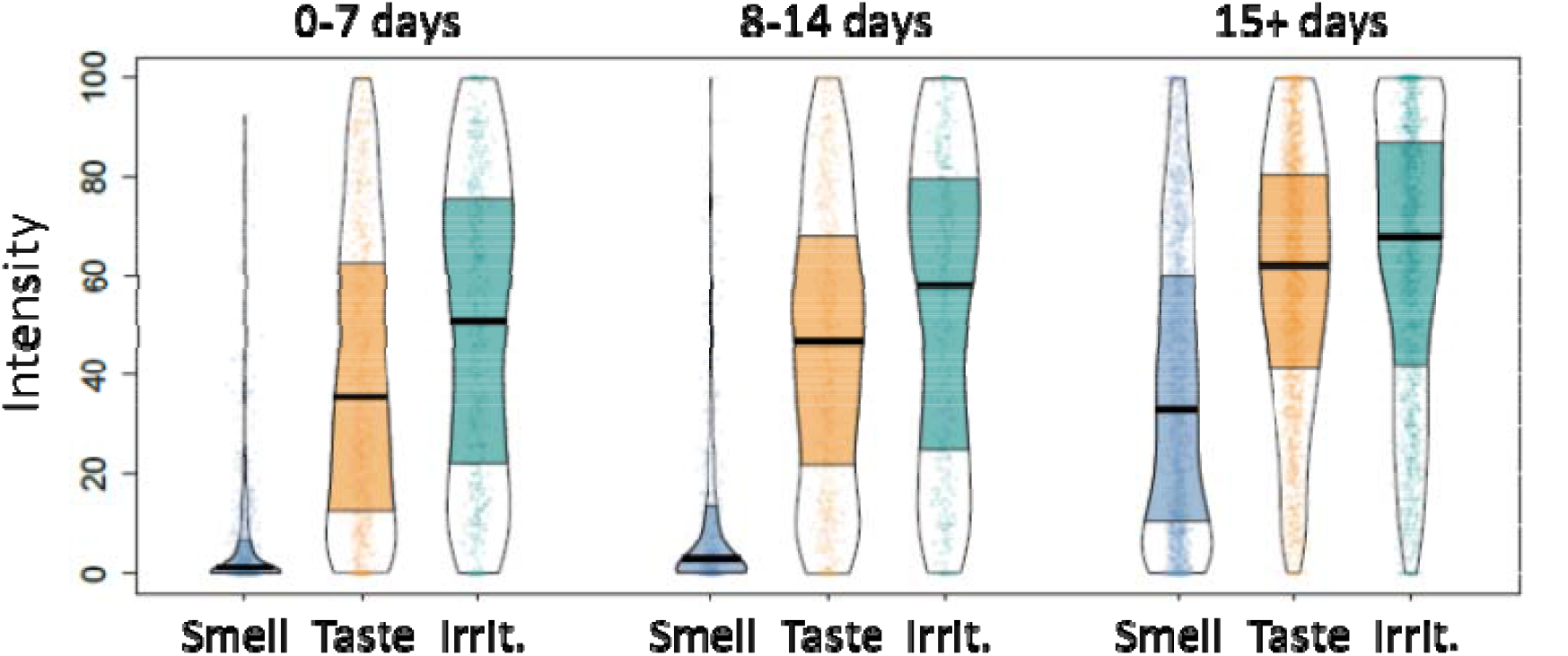
Perceived intensities for smell, taste, and irritation in participants who completed the survey within the first 7 days (n=737), 8-14 days (n=597), and >15 days (n=1,831) of the onset of COVID-19 illness. Points represent individual subject data (jittered horizontally), the center horizontal bars depict medians, the shapes reflect the density of the distribution, and the colored areas show interquartile range. Note that only participants who reported a COVID diagnosis date are presented here.

**Supplementary Figure 3.**
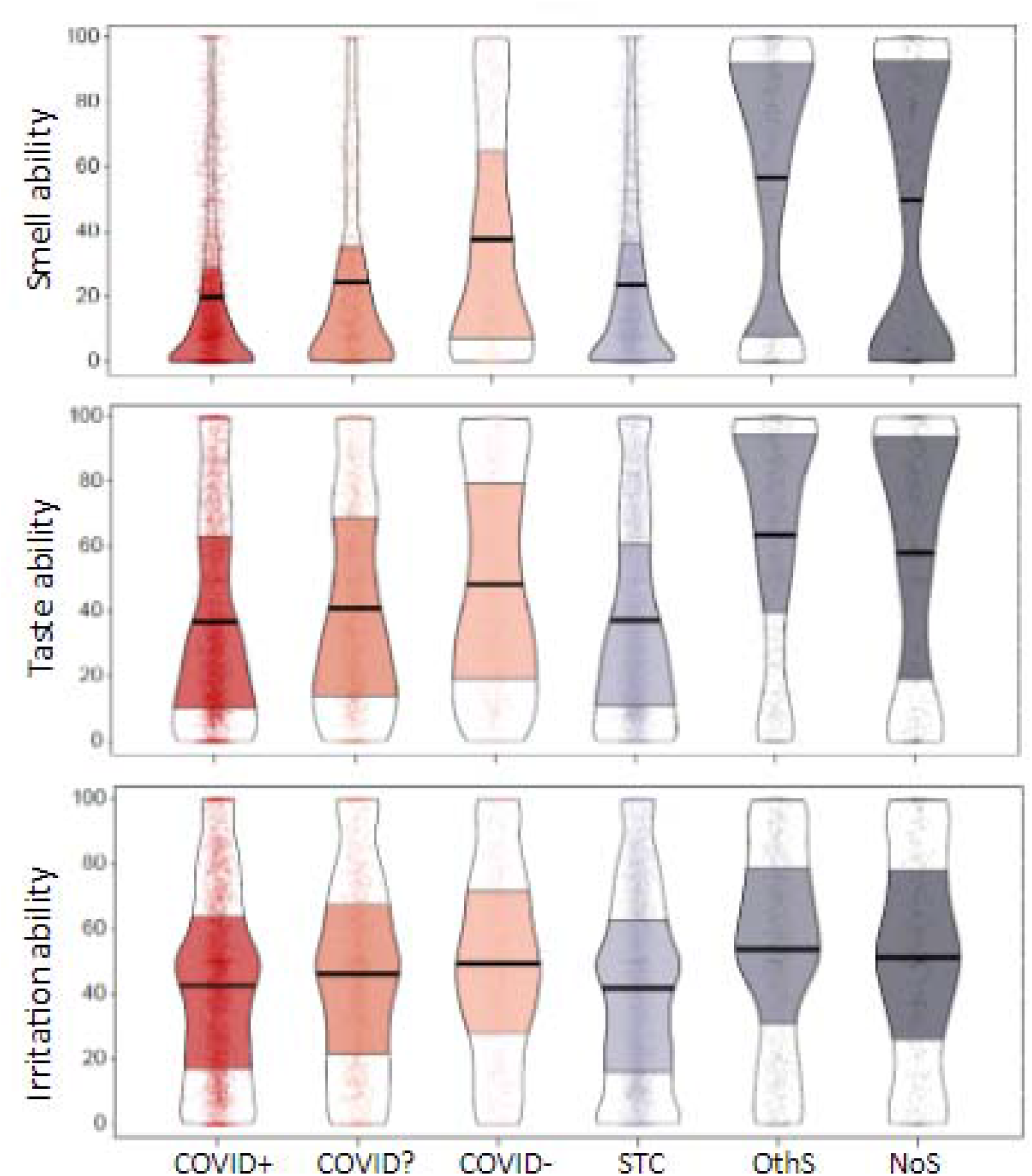
Self-reported ability to taste, smell, and experience irritation for six groups. Participants are grouped according to COVID diagnosis or symptoms (from left to right) into COVID positive (COVID+; N=3,275), participants who were awaiting COVID test results (COVID?; N=1,224), COVID negative (COVID-; N=579),, those who reported sudden smell/taste changes (STC; N=4,271), and those with other symptoms excluding smell or taste loss (OthS; N=802), and no symptoms (NoS; N=396). They indicated perceived intensity of smell, taste, and irritating stimuli using a visual analog scale (0-100). Points represent individual subject data (jittered horizontally), the center horizontal bars depict the medians, the shapes reflect the density of the distribution, and the colored areas show interquartile range. See **Figure 2** for the corresponding perceived chemosensory intensities.

**Supplementary Figure 4.**
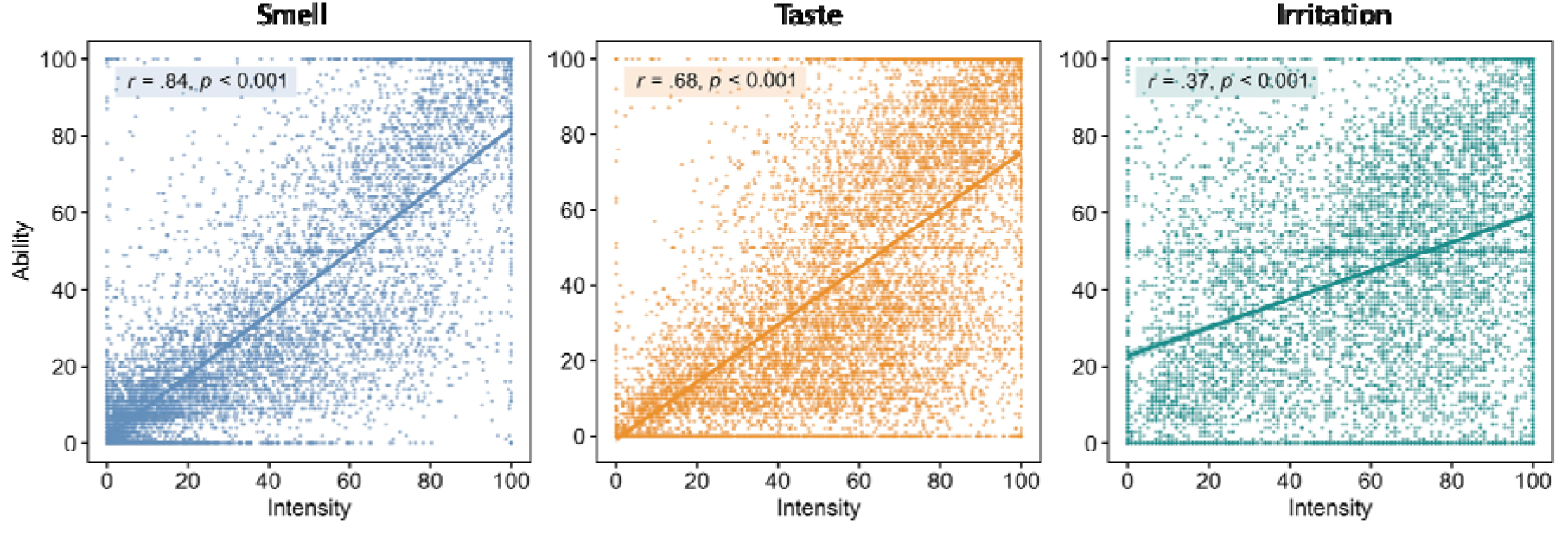
Correlations between chemosensory intensity perception in the home test and the corresponding self-reported chemosensory ability. Dots represent individual subject data, the lines are for linear regression between the intensity and ability ratings for each chemosense, smell, taste, and oral irritation.

## Supplementary Tables

**Supplementary Table 1.** Foods and household items presented in the hometest.

| modality | category | items |
| --- | --- | --- |
| Smell | Cosmetics | skin lotion; laundry detergent; shampoo or conditioner; perfume; scented soap; oil for body or hair care; sunscreen |
|  | Fruits | pineapple; apple; apricot; banana; strawberry; green or spring onion; melon or watermelon; bell pepper (not spicy); peach; orange; cucumber; tomato; lemon |
|  | Spices | asafetida; oyster or fish sauce; fennel seeds; garam masala; ginger; kaffir lime; cardamom; coconut oil; coriander seeds; cumin; caraway; turmeric; lavender; nutmeg; cloves; oregano; panch phoron; rose oil; rosemary; sesame oil; soy sauce; cinnamon |
|  | Other | freshly roasted toast or bread; crisps or chips; canned fish; peanut butter; smoked food (lamb or fish or other); coffee; cheese; cat food; biscuits or cookies; dried seafood; chocolate; chocolate spread (for example, Nutella); shoe cream; tea; joss-stick or frankincense; cigarette butts or ashtray; burnt matches |
| Taste | Sweet | sugar or other sweetener |
|  | Sour | vinegar (any kind but balsamic) or lemon or lime juice |
|  | Salty | salt |
|  | Bitter | coffee (beans, grounds, instant powder) or black or green tea leaves |
| Irritation | Oral | hot mustard; horseradish; wasabi; chili pepper (fresh / ground / dried); ginger (fresh / ground); hot sauce; carbonated water (normal sip); mint candy |
|  | Nasal | vinegar; mustard; horseradish; Vicks Vaporub® or Camphor or Eucalyptus oil |

**Supplementary Table 2.** Subject characteristics of the six diagnostic groups.

|  | Respiratory illness |  |  | No respiratory illness |  |  |  |
| --- | --- | --- | --- | --- | --- | --- | --- |
|  | COVID+<br>*n* = 3,356 | COVID?<br>*n* = 1,267 | COVID-<br>*n* = 602 | STS<br>*n* = 4,445 | OthS<br>*n* = 832 | NoS<br>*n* = 416 | Significance<sup>1</sup> |
| Gender, *n* |  |  |  |  |  |  | *** |
| Women | 2604 | 964 | 443 | 3091 | 526 | 236 |  |
| Men | 740 | 287 | 153 | 1287 | 290 | 174 |  |
| Other | 2 | 3 | 1 | 14 | 5 | 2 |  |
| Not shared | 10 | 13 | 5 | 53 | 11 | 4 |  |
| Age, yrs<br>(mean±SD) | 39.46±12.83 | 37.15±12.61 | 41.77±14.05 | 38.42±13.82 | 41.39±15.43 | 44.81±16.83 | *** |
| SES, *n* |  |  |  |  |  |  | *** |
| Upper | 217 | 53 | 34 | 210 | 42 | 31 |  |
| Upper mid | 1689 | 505 | 241 | 1927 | 361 | 196 |  |
| Lower mid | 924 | 431 | 214 | 1318 | 247 | 103 |  |
| Lower | 98 | 94 | 38 | 191 | 38 | 12 |  |
| Not shared | 428 | 184 | 75 | 799 | 144 | 74 |  |
| Smoking, *n* |  |  |  |  |  |  | *** |
| Current | 424 | 191 | 98 | 751 | 136 | 50 |  |
| Former | 737 | 239 | 123 | 886 | 145 | 91 |  |
| Never | 2,150 | 813 | 371 | 2,751 | 540 | 263 |  |
| Not shared | 45 | 24 | 10 | 57 | 11 | 12 |  |

<sup>1</sup> Significance of the ANOVA or Chi-squared test with n.s. (=not significant) for *p*-value > 0.05 and
\*\*\* for *p*-value ≤ 0.001

**Supplementary Table 3.** Pairwise comparisons the six diagnostic groups for perceived intensity and reported ability.

| Tukey contrasts | ability |  |  | intensity |  |  |
| --- | --- | --- | --- | --- | --- | --- |
|  | smell | taste | irritation | smell | taste | irritation |
| COVID+ vs. COVID- | < 0.001 | < 0.001 | < 0.001 | < 0.001 | < 0.001 | 0.006 |
| COVID+ vs. NoS | < 0.001 | < 0.001 | < 0.001 | < 0.001 | < 0.001 | < 0.001 |
| COVID+ vs. OthS | < 0.001 | < 0.001 | < 0.001 | < 0.001 | < 0.001 | < 0.001 |
| COVID+ vs. STC | < 0.001 | 0.997 | 0.687 | < 0.001 | 0.998 | 0.984 |
| COVID+ vs. COVID? | < 0.001 | < 0.001 | 0.003 | 0.009 | 0.189 | 0.999 |
| COVID- vs. NoS | < 0.001 | < 0.001 | 0.944 | 0.003 | 0.006 | 0.023 |
| COVID- vs. OthS | < 0.001 | < 0.001 | 0.056 | < 0.001 | < 0.001 | < 0.001 |
| COVID- vs. STC | < 0.001 | < 0.001 | < 0.001 | < 0.001 | < 0.001 | 0.001 |
| COVID- vs. COVID? | < 0.001 | < 0.001 | 0.222 | < 0.001 | 0.002 | 0.012 |
| NoS vs. OthS | < 0.001 | 0.027 | 0.621 | < 0.001 | 0.545 | 0.700 |
| NoS vs. STC | < 0.001 | < 0.001 | < 0.001 | < 0.001 | < 0.001 | < 0.001 |
| NoS vs. COVID? | < 0.001 | < 0.001 | 0.037 | < 0.001 | < 0.001 | < 0.001 |
| OthS vs. STC | < 0.001 | < 0.001 | < 0.001 | < 0.001 | < 0.001 | < 0.001 |
| OthS vs. COVID? | < 0.001 | < 0.001 | < 0.001 | < 0.001 | < 0.001 | < 0.001 |
| STC vs. COVID? | 0.968 | 0.001 | < 0.001 | 0.999 | 0.077 | 0.999 |

**Supplementary Table 4.** Posthoc tests for perceived intensities for smell, taste, and irritation in participants who completed the survey within the first 7 days (n=737), 8-14 days (n=597), and >15 days (n=1,831) of the onset of COVID-19 illness.

| Mean ± SD |  |  |  |  |  |
| --- | --- | --- | --- | --- | --- |
| Group | 0-7 days<br>(n=737) | 8-14 days<br>(n = 597) | >15 days<br>(n = 1831) | p-value<br>(ANOVA) | Eta.[95% CI] |
| Smell intensity | 6.7 ± 13.1<sup>a</sup> | 11.2 ± 18.3<sup>b</sup> | 37.3 ± 29.2<sup>c</sup> | <2e-16 | 0.25 [0.18, 0.28] |
| Taste intensity | 39.6 ± 29.3<sup>a</sup> | 46.1 ± 28.6<sup>b</sup> | 59.4 ± 26.2<sup>c</sup> | <2e-16 | 0.09 [0.07, 0.11] |
| Oral-irritation intensity | 50.0 ± 30.7<sup>a</sup> | 53.4 ± 31.1<sup>a</sup> | 63.7 ± 28.2<sup>b</sup> | <2e-16 | 0.04 [0.03, 0.06] |

**Supplementary Table 5.**
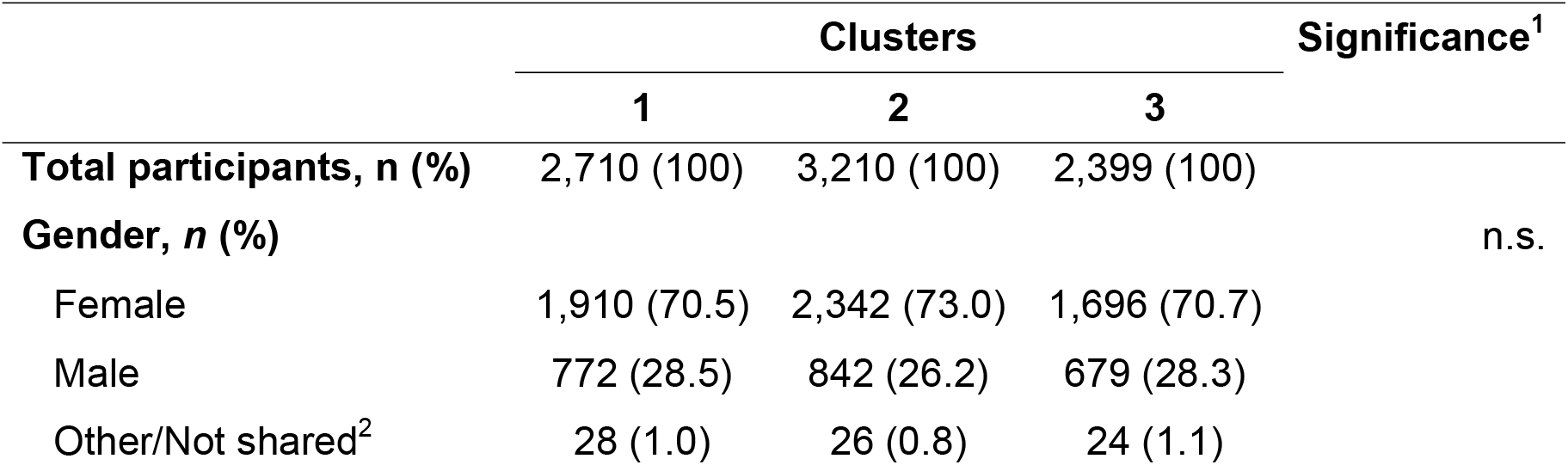
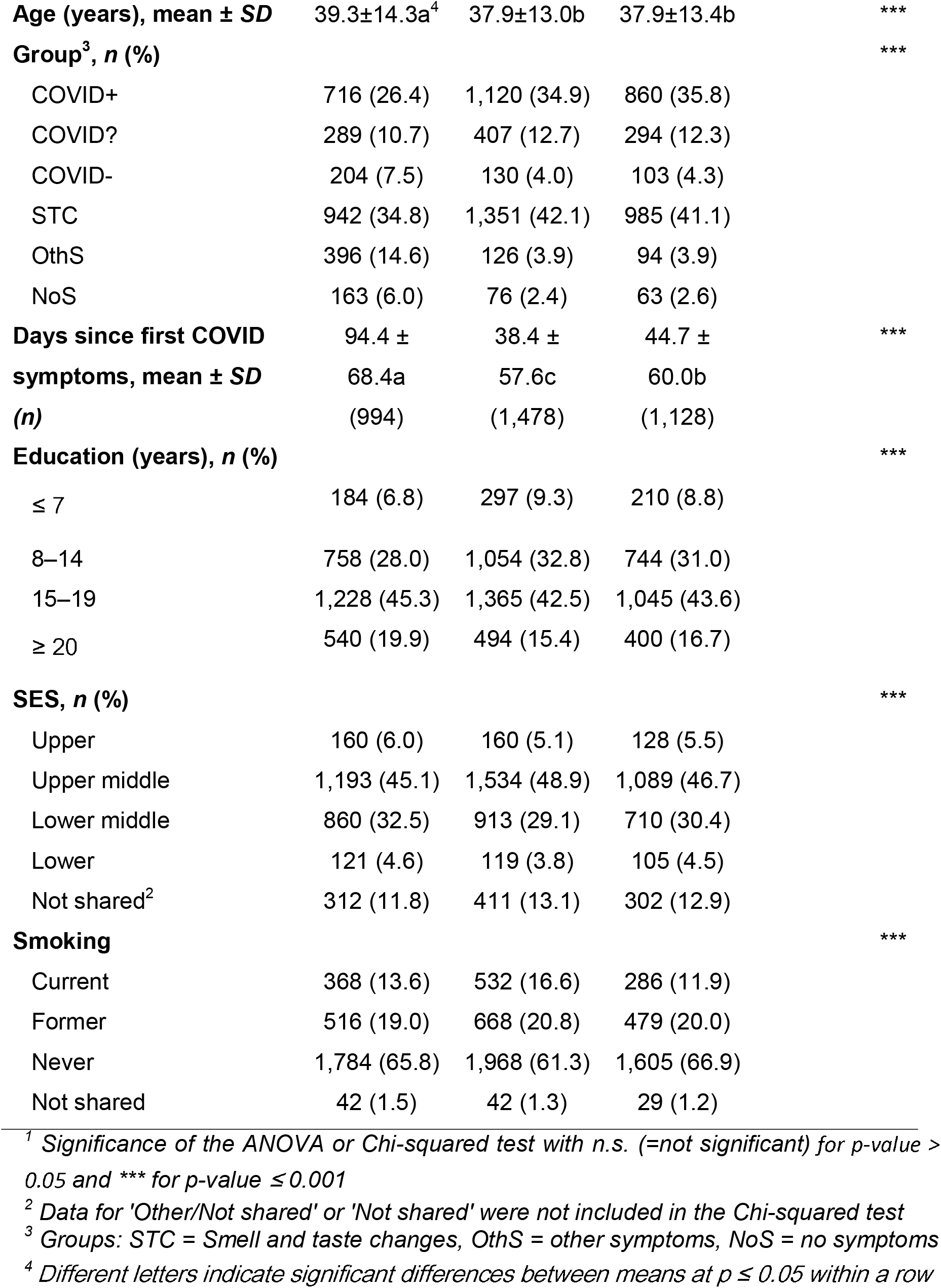
Subject characteristics for 3 clusters identified by the Agglomerative Hierarchical Clustering (AHC) on perceived intensities.

